# *MUC1* and *MUC4* expression are inversely correlated and trigger immunological response and transport pathways in gliomas

**DOI:** 10.1101/2024.06.28.24309676

**Authors:** Gabriel Cardoso Machado, Valéria Pereira Ferrer

## Abstract

Diffuse gliomas are tumors that arise from glial or glial progenitor cells. They are currently classified as astrocytoma isocitrate dehydrogenase (IDH)-mutant or oligodendroglioma IDH-mutant, and chromosome arms 1p/19q-codeleted, both slower-growing tumors, or glioblastoma (GBM), a more aggressive tumor. Despite advances in the diagnosis and treatment of gliomas, the median survival time after diagnosis of GBM remains low, approximately 15 months, with a 5-year overall survival rate of only 6.8%. Therefore, new biomarkers and therapy targets that could support better prognosis of these tumors would be of great value. MUC1 and MUC4, membrane-bound mucins, has been identified as a potential biomarker in several tumors. However, the role of these mucins in adult gliomas has not yet been well explored. Here, we show for the first time, in a retrospective study and by *in silico* analysis, the relevance and correlation of these genes in adult gliomas. Analysis of adult diffuse glioma patient cohorts revealed differential methylation and expression patterns of MUC1 and MUC4 across GBM and non-GBM subtypes. GBM patients exhibited decreased MUC1 methylation and elevated expression (r-0.25, p < 0.0001) whereas increased MUC4 methylation and its lower expression (r-0.13, p = 0.1344). Conversely, in non-GBM patients, MUC1 showed higher methylation levels and low expression (r-0.27, p < 0.0001) whereas MUC4 showed lower methylation levels and high expression (r-0.32, p < 0.0001). The expression levels of these genes influenced overall survival (OS) in gliomas patients (p = 0.0344), with high MUC1 and low MUC4 expression associated with worse OS. MUC1 and MUC4 correlated with MUC20 in both GBM (r = 0.54) and non-GBM (r = 0.53) patients (p < 0.0001). Functional enrichment analysis revealed distinct biological roles for co-expressed genes with MUC1 involvement in innate immunity, antigen processing, pro-inflammatory responses in both non-GBM and GBM cohorts, and integrin-based signaling pathways in GBM patients. MUC4 co-expressed genes were involved in ion transport in GBM patients. Using molecular docking, we observed that MUC1 has domains that are physically capable of interacting with immune response-related proteins such as Receptor for Advanced Glycation End-products (RAGE), Major Histocompatibility Complex II (MHCII), and extracellular matrix receptor integrin alpha 2 (ITGA2). These findings shed light on the molecular mechanisms underlying glioma progression and highlight MUC1 and MUC4 as potential prognostic markers and therapeutic targets in glioma management.

## 1. Introduction

Diffuse gliomas constitute a group of neuroepithelial tumors originating from neuroglia or their precursor cells [1]. These tumors are characterized by high biological and morphological heterogeneity, making them the most prevalent malignant primary tumors of the brain [2]. Historically, gliomas were classified based on histopathological features. However, with the advent of cancer multiomics, molecular biomarkers have been identified, enabling the delineation of more precisely defined and biologically uniform disease entities [3,4].

For adult-type diffuse gliomas, mutations in isocitrate dehydrogenase (*IDH*) 1/2 and codeletion of the 1p and 19q chromosome arms have been identified as significant prognostic factors, forming the foundation for glioma classification in the fourth revised edition of the WHO classification of CNS tumors in 2016 [5,6]. This classification system also considers tumor grade (1-4), reflecting the aggressive nature of glioma [7]. The most recent update, released in 2021 (WHO2021), further refines the classification of adult-type diffuse glioma into astrocytoma *IDH*-mutant (grade 2-4), oligodendroglioma *IDH*-mutant and 1p/19q-codeleted (grade 2-3), and glioblastoma (GBM), *IDH*-wildtype (grade 4) [8]. In this study, we collectively refer to astrocytoma and oligodendroglioma as non-GBM gliomas.

GBM is the most common and aggressive primary malignant brain tumor, representing 59.2% of all gliomas and 50.1% of all malignant tumors of the brain [9]. Even with the gold standard treatment for GBM, which includes maximal total resection and DNA-damaging chemo-/radiotherapy [10], the disease remains incurable, with a global OS of 14 months [11]. GBM malignancy is linked to the potential of tumor cells to restructure the extracellular matrix (ECM), ensuring invasiveness in the brain parenchyma [12]. Therefore, even after tumor resection, the infiltration of residual cells triggers tumor relapse, underscoring the urgent need for the development of new biomarkers and therapies for GBM treatment [13].

Mucins constitute a family of heavily glycosylated proteins with high molecular weights that are renowned for their roles in lubrication and epithelial protection [14]. In *Homo sapiens*, at least 20 genes have been identified, giving rise to proteins classified as membrane-bound mucins or secreted mucins [15]. The general mucin structure is characterized by the presence of domains rich in proline, threonine and serine (PTS) residues that occur in tandem and undergo extensive O-glycosylation [14]. Specifically, those tethered to the cell membrane, such as MUC1 and MUC4, not only feature transmembrane domains but also possess a cytoplasmic tail that can undergo phosphorylation, playing a role in signal transduction [16].

MUC1 (aa 1255) and MUC4 (aa 2169) are type I transmembrane proteins with heavily glycosylated extracellular domains extending 200-500 nm and >2 μm from the cell surface, respectively [16–18]. In healthy tissues, these mucins are expressed by epithelial cells, providing a barrier to pathogens and regulating inflammatory responses [19,20]. In several cancers, MUC1 and/or MUC4 are overexpressed and abnormally glycosylated [21–24]. Very little is known about the role of these two mucins in gliomas. MUC1 contributes to resistance, cell cycle regulation and malignancy [25,26], and MUC4 contributes to invasion, proliferation and malignancy [27,28].

In this study, for the first time, we investigated the possible connection between MUC1 and MUC4 in gliomas. Using clinical data from glioma patients and *in silico* analysis, we assessed the methylation and expression levels of *MUC1* and *MUC4* in glioma patients according to different parameters (subtypes, grades and WHO 2021 classification) and identified correlations between *MUC1*/*MUC4* methylation and expression levels in both GBM and non-GBM glioma cohorts. We demonstrated that adult patients with gliomas with high *MUC1* and low *MUC4* expression had worse OS. Furthermore, we found that genes coexpressed with *MUC1* and *MUC4* are related to both innate and adaptative immune responses, extracellular matrix-integrin-based signaling and ion transport. We also observed that the SEA (sea urchin sperm protein, enterokinase, agrin) domain of MUC1 can physically interact with proteins related to the immune response and cell migration.

## 2. Methods

### 2.1 *MUC1* and *MUC4* methylation analysis in adult diffuse gliomas

Clinical data regarding gene methylation in glioma patients were first obtained from the Chinese Glioma Genome Atlas (CGGA, http://www.cgga.org.cn/) [29]. Specifically, we utilized data from the “methyl_159” study, which included 151 glioma patients (both primary and recurrent) and 8 control individuals, to assess the methylation of 36,326 genes. In our study, we focused on *MUC1* and *MUC4* data. Methylation quantification was scaled from 0 (hypomethylated) to 1 (hypermethylated) [30].

According to the cohort’s classification, we considered histology, *IDH* mutation status and 1p/19q codeletion status. In the non-GBM cohort, patients with wild-type *IDH* status were excluded, while in the GBM cohort, patients with mutated *IDH* status were excluded. Patients whose histology was not currently recognized in the WHO2021 classification were also excluded. Other molecular parameters based on the latest glioma classification (WHO2021) were also considered for the generation of the cohorts.

### 2.2 *MUC1* and *MUC4* expression in adult diffuse gliomas

Gene expression and clinical data were first obtained from the CGGA (http://www.cgga.org.cn/) [29]. For expression analysis, we utilized data from the “mRNAseq_693” [31–33] and “mRNAseq_325” [31,34,35] studies, which included a total of 1,019 patients with gliomas. Data on gene expression levels, in fragments per kilobase of transcript per million mapped reads (FPKM), were obtained for 23,987 and 24,326 genes, respectively. In this study, we focused on the expression of the *MUC1* and *MUC4* genes. Cohort classification followed the same criteria mentioned earlier.

### 2.3 Correlation analysis between *MUC1* and *MUC4* methylation and expression

The correlation analysis between methylation and expression was carried out using data from glioma patients available on cBioPortal (https://www.cbioportal.org/) [36–38]. This platform was chosen due to its ability to provide individual assessments of methylation and expression for the same patient and to increase the number of patients analyzed.

The methylation and expression data were accessed on cBioPortal using the Firehose Legacy Study from The Cancer Genome Atlas (TCGA) to construct cohorts for both GBM (https://gdac.broadinstitute.org/runs/stddata 2016_01_28/data/GBM/20160128/) and non-GBM (https://gdac.broadinstitute.org/runs/stddata2016_01_28/data/LGG/20160128/) patients. For the non-GBM cohort, methylation data from the HumanMethylation450 (HM450) platform comprising 530 patients were utilized. For the glioma GBM cohort, data from 428 patients from both the Human Methylation 27 (HM27) and HM450 platforms were used. For both cohorts, the methylation data for the *MUC1* and *MUC4* genes were considered, ranging from 0 (hypomethylated) to 1 (hypermethylated). We utilized mRNA expression z score (log RNA Seq V2 RSEM) data from the same set of patients described for the methylation data. Correlation analysis between the methylation and expression of the *MUC1* and *MUC4* genes in glioma patient data was conducted using the nonparametric Spearman correlation coefficient, which ranges from -1 (inverse correlation) to +1 (perfect correlation).

### 2.4 Overall survival analysis

The OS of GBM patients (N=153) with high or low expression of *MUC1* and *MUC4* was assessed using data from The Human Protein Atlas (https://www.proteinatlas.org/) [39, 40] . The database provides a cutoff that represents the optimal expression value (FPKM) yielding the maximal difference in survival between the two groups. The cohorts were subdivided into patients with high or low *MUC1* or *MUC4* expression (cutoff = 3.39 and 0.07, respectively).

Survival comparisons between GBM patients with high or low *MUC1* and *MUC4* expression were conducted using the Kaplan‒Meier method and the log-rank (Mantel– Cox) test. The median survival time was calculated as the shortest survival time for which the survival function was equal to or less than 50%.

### 2.5 Coexpression and enrichment analysis

Data regarding genes that are individually coexpressed with *MUC1* and *MUC4* in gliomas were accessed via cBioPortal (https://www.cbioportal.org/) using the PanCancer Atlas study from TCGA for both GBM and non-GBM [41–50] cohorts. In the coexpression tab, we downloaded lists of genes positively and negatively correlated with *MUC4* for the non-GBM cohort and *MUC1* for the GBM cohort. Genes with Spearman’s coefficient above 0.4 were selected for enrichment analysis in both cohorts.

Enrichment analysis was conducted using a database for annotation, visualization and integrated discovery (DAVID) from the National Institute of Health (NIH, https://david.ncifcrf.gov/) [51,52]. Biological processes from Gene Ontology and functional annotations were used for clustering. The group enrichment score (ES), the geometric mean (on a -log scale) of a member’s p value in a corresponding annotation cluster, was used to rank its biological significance.

### 2.6 Protein‒protein docking analysis

To predict potential physical interactions between the MUC1 SEA domain and components of innate and adaptive immune responses, as well as integrin signaling, based on our enrichment results, molecular docking experiments were performed using the ClusPro 2.0 server (https://cluspro.bu.edu/home.php). The ClusPro server is based on PIPER, a fast Fourier transform-based rigid docking program that attempts to find the native site under the assumption that it will have a wide free-energy attractor with the largest number of results. ClusPro provides energy scores from the PIPER [53–56].

MUC1 and galectin-3/galectin-3-binding protein (Gal-3-BP) are known to interact [57,58], and this interaction was used as a control. To explore the interaction of MUC1 with proteins from related pathways, we selected the RAGE protein, an important receptor that orchestrates the innate immune response; the MHCII protein, which is crucial for antigen presentation in the adaptive immune response; and integrin alpha2, which is involved in integrin signaling. The following crystal structures of these human proteins were obtained from the Protein Data Bank (PDB, https://www.rcsb.org/?ref=nav_home): MUC1 SEA domain (6BSC), HLA-DP (7ZFR), RAGE VCI domain (7LMW), domain I from integrin alpha2 beta1 (1AOX), and Gal-3-BP (6GFB). Regarding MUC4, there was no resolved crystal structure available in the database. Therefore, we were unable to perform the analysis.

### 2.7 Statistical analysis

The statistical analysis and graph generation were performed using GraphPad Prism (version 9). For group comparisons, we initially conducted normality and log normality tests (D’Agostino and Pearson tests). After confirming that the data did not follow a normal distribution, the Mann‒Whitney test was used for independent samples, and the Wilcoxon rank test was used for matched-pairs samples. The Kruskal‒Wallis test was utilized for comparing three or more groups, and Dunn’s multiple comparisons test was used when applicable.

OS analyses, utilizing the Kaplan‒Meier method and the log-rank (Mantel‒Cox) test, and Spearman’s correlation test were also performed in GraphPad Prism.

## 3. Results

### 3.1 *MUC1* expression is increased in GBM, whereas *MUC4* expression is increased in astrocytomas and oligodendrogliomas

The methylation profiles of the *MUC1* and *MUC4* genes were assessed using data from adult diffuse glioma patients in the CGGA. Patients were initially classified into GBM (N=29) and non-GBM (N=70) glioma cohorts. *MUC1* methylation was significantly greater (p <0.0001) in non-GBM patients (median =0.21) than in GBM patients (median = 0.18) (Fig. 1A). Conversely, *MUC4* methylation was significantly greater (p = 0.0006) in GBM patients (median =0.86) than in non-GBM patients (median =0.74) (Fig. 1B).

**Fig. 1.**
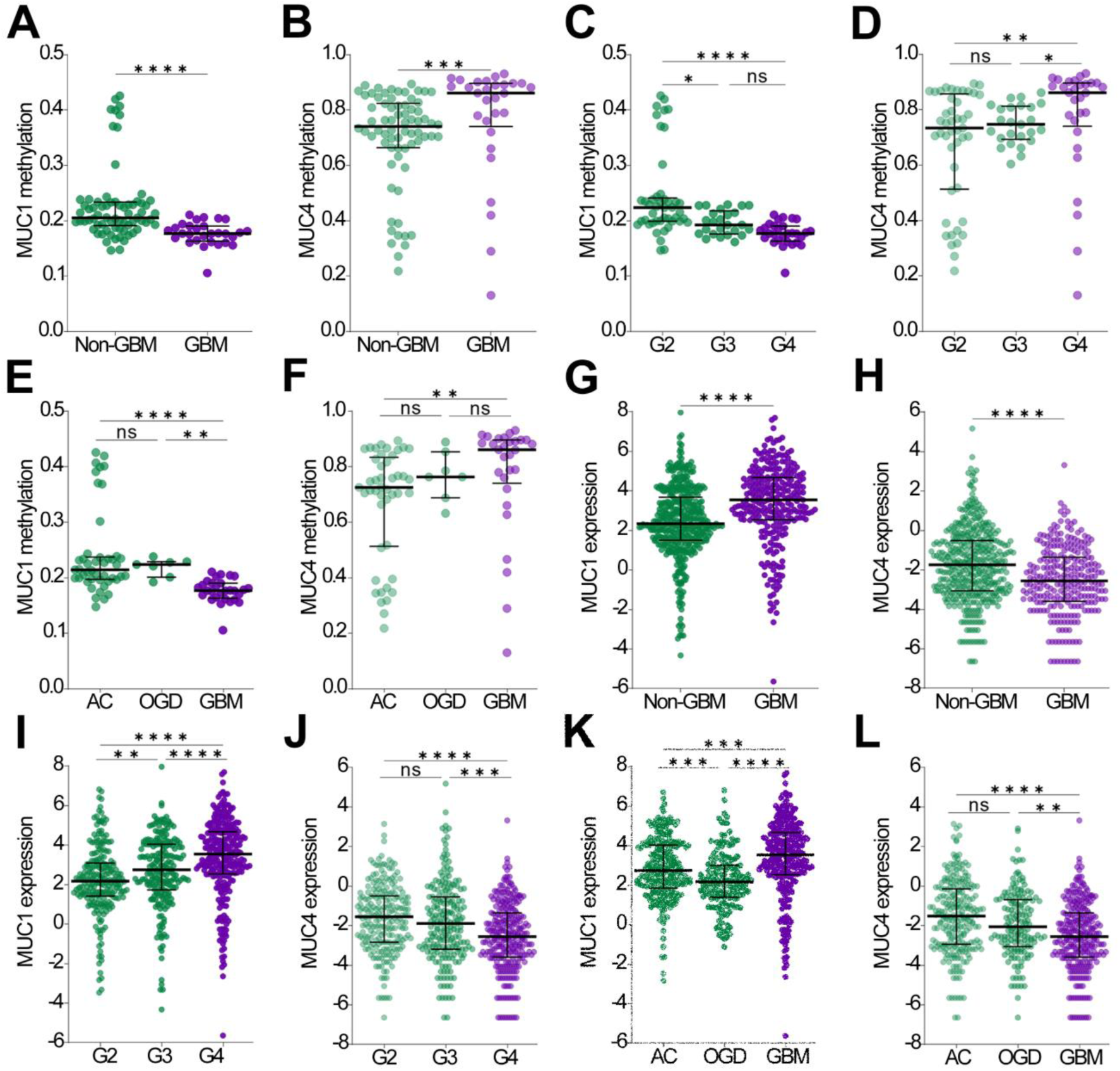
*MUC1* and *MUC4* methylation and expression are distinctly associated with tumor subtype and grade. *MUC1* and *MUC4* methylation (N = 99) or expression (N = 709) were analyzed using CGGA data. Glioma samples were categorized into non-GBM versus GBM groups (A, B, G and H), grade 2, 3 or 4 glioma groups (C, D, I and J) and AC versus OGD or GBM groups (E, F, K and L). In GBM patients, *MUC1* was less methylated and more highly expressed than in non-GBM patients. *MUC1* methylation decreased with increasing tumor grade and increased expression. For *MUC4*, the opposite trend was observed. D’Agostino-Pearson and Mann‒Whitney tests were performed for statistical analysis (p > 0.05, not significant; *p < 0.05; **p < 0.01; *** p < 0.001; **** p < 0.0001). *MUC1* data are represented in dark colors, and *MUC4* data are represented in light colors. Green represents the non-GBM or lower-grade glioma group, and purple represents the GBM group.

Further analysis revealed significant differences in *MUC1* and *MUC4* methylation between Grade 2 (G2) (N =45), Grade 3 (G3) (N = 25) and Grade 4 (G4) (N = 29) patients (p < 0.0001 and p =0.0035, respectively). Over the tumor grade, *MUC1* methylation decreased (G2, median = 0.22; G3, median = 0.19; G4, median = 0.18), with G2 showing a significant increase compared to G3 (p = 0.0134) and G4 (p <0.0001) (Fig. 1C). In contrast, MUC4 methylation increased with increasing tumor grade in the G4 subgroup (median = 0.86) compared to the G3 subgroup (median = 0.75; p = 0.0255) and G2 subgroup (median = 0.73, p = 0.0046) (Fig. 1D).

The methylation of *MUC1* and *MUC4* was also assessed following the WHO2021 glioma classification, categorizing samples into astrocytoma *IDH*-mutated (AC) (N=45); oligodendroglioma, IDH-mutant and 1p/19q-codeleted (OGD); and GBM, *IDH* wild type (N= 29). Significant differences in *MUC1* and *MUC4* methylation were observed among the cohorts (p < 0.0001 and p = 0.0035, respectively). Compared with OGD (median = 0.22; p = 0.0032) and AC (median = 0.21; p < 0.0001), GBM exhibited lower *MUC1* methylation (median = 0.18) (Fig. 1E). GBMs had greater *MUC4* methylation (median = 0.86) than did ACs (median = 0.73; p = 0.0019), with no significant difference observed compared to that of OGDs (median = 0.76; p = 0.7987) (Fig. 1F).

The mRNA expression of *MUC1* and *MUC4* was evaluated using a different dataset from the CGGA. After classifying patients into GBM (N = 275) and non-GBM (N = 434) glioma cohorts, a significant (p <0.0001) increase in MUC1 expression was observed in GBM patients (median = 3.54) compared to non-GBM patients (median = 2.33) (Fig. 1G). Conversely, *MUC4* expression was significantly greater in the non-GBM cohort (median -1.737) than in the GBM cohort (median = -2.556) (Fig. 1H).

Considering tumor grade (G2, N=223; G3, N=211; G4, N=275), a significant (p <0.0001 for both cohorts) variation in *MUC1* and *MUC4* expression was observed. MUC1 mRNA levels increased with tumor malignancy, particularly in G4 patients (median = 3.54), compared to G3 patients (median = 2.75) and G2 patients (median = 2.17) (p <0.0001 for both cohorts) (Fig. 1I). A significant difference was also observed between G2 and G3 patients (p = 0.0047) (Fig. 1I). Accordingly, *MUC4* expression decreased with increasing tumor grade, especially in G4 (median -2.556) patients compared to G3 (median -1.889; p = 0.0005) and G2 (median -1.556; p <0.0001) patients (Fig. 1J).

*MUC1* and *MUC4* expression was also assessed according to the WHO2021 glioma classification. Both mRNA expression levels varied significantly among the AC (N=244), OGD (N=158) and GBM (N=275) patient cohorts (p <0.0001 for both). *MUC1* expression was significantly greater in GBM patients (median = 3.54) than in patients with OGD (median = 2.17, p <0.0001) or AC (median = 2.75; p=0.0002), with a significant difference between the ODG and AC patient cohorts (p = 0.0002) (Figure 2K). However, the MUC4 mRNA level was significantly lower in GBM patients (median - 2.556) than in OGD patients (median -2.059; p = 0.0032) and AC patients (median -1.515; p <0.0001) (Fig. 1L).

**Fig. 2.**
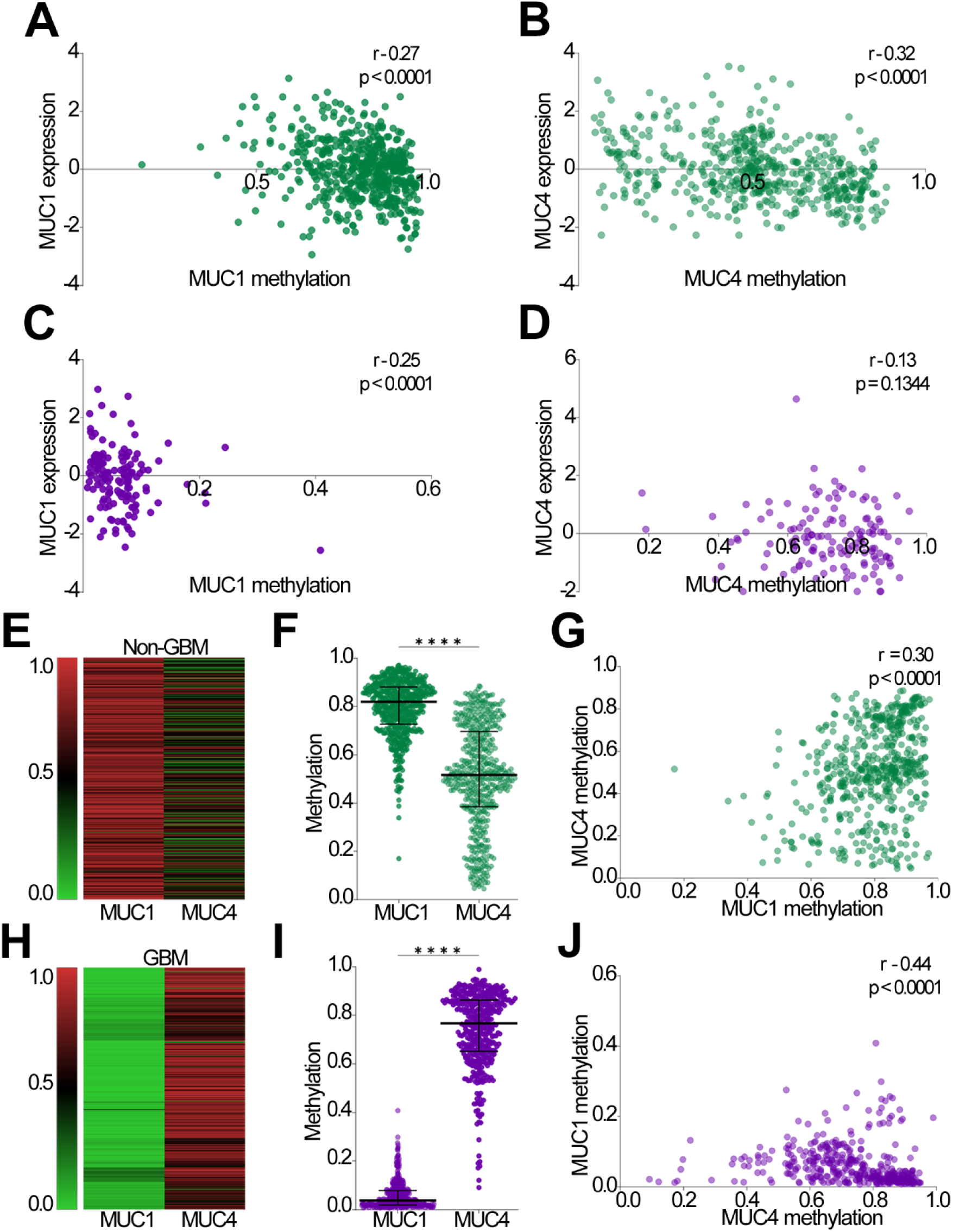
Methylation of *MUC1* and *MUC4* is correlated in glioma patients and suppresses their respective expression. The methylation and expression of *MUC1* and *MUC4* were analyzed in glioma patients. Significant correlations were observed in the non-GBM (A-B, N = 530) and GBM (C-D, N = 137) groups, with the exception of *MUC4* in GBM. In non-GBM patients, *MUC1* showed higher methylation levels than *MUC4* (E-F), and these levels were positively correlated (G), indicating increased methylation of both genes in lower-grade gliomas. Conversely, in GBM patients, *MUC1* methylation was lower than *MUC4* methylation (H-I), with a negative correlation between the genes (J), indicating that increased *MUC4* methylation leads to decreased *MUC1* methylation. Statistical analyses were performed using the D’Agostino-Pearson test, Wilcoxon rank test for matched-pairs samples, and Spearman correlation analysis (**** p < 0.0001). Green represents the non-GBM or lower-grade glioma group, and purple represents the GBM group.

Therefore, an apparent inverse correlation between methylation and *MUC1* and *MUC4* expression was observed in all the analyses. *MUC1* was more highly expressed in GBM, whereas *MUC4* expression was greater in AC and OGD.

### 3.2 *MUC1* and *MUC4* methylation and expression are distinctly correlated in non-GBM and GBM cohorts

To confirm the previous results, where we identified the methylation of *MUC1* and *MUC4* in a group of glioma patients and the expression pattern in another cohort of glioma patients, we investigated the correlation between *MUC1* and MUC4 methylation and expression by assessing these parameters in the same glioma cohort.

To validate the impact of methylation level alterations on gene expression, we investigated the correlation between methylation and expression data in non-GBM (N = 530) and GBM (N = 137) cohorts. For the non-GBM group, both genes demonstrated a significant (p <0.0001) negative correlation (MUC1, r - 0.27; MUC4, r - 0.32), indicating that increasing methylation of these genes decreases their expression (Fig. 2A-B). In the GBM cohort, a similar pattern was observed for MUC1 (r = - 0.25, p = 0.0027) (Fig. 2C). However, for MUC4 in GBM, these parameters were not significantly correlated (r - 0.13, p = 0.1344) (Fig. 2D).

In the non-GBM cohort (n=530), MUC1 methylation was significantly greater (median = 0.82, p < 0.0001) than MUC4 methylation was (median = 0.52) (Fig. 2E-F). A positive correlation (r = 0.30; p < 0.0001) was observed, indicating that increasing MUC1 methylation corresponds to an increase in MUC4 methylation (Fig. 2G).

In turn, in the GBM cohort (N = 428), there was a significant increase in MUC4 methylation (median = 0.77, p <0.0001) compared to that in the MUC1 cohort (median = 0.04) (Fig. 2H-I). Correlation analysis revealed a negative correlation (r=-0.44; p<0.0001), suggesting that an increase in MUC4 methylation corresponds to a decrease in MUC1 methylation (Fig. 2J).

### 3.3 *MUC1* and *MUC4* expression impact OS and are correlated with MUC20 in both GBM patients and non-GBM patients

To confirm the correlation between *MUC1* and *MUC4* gene expression, we investigated whether their expression levels impact glioma patient survival (N = 153). We observed significantly shorter survival in patients with high MUC1 expression (OS= 10.82; p = 0.0143) than in patients with low *MUC1* expression (OS= 14.93; N= 103). Conversely, for *MUC4* expression, patients with low *MUC4* expression (N=118) had shorter survival (OS= 12.76; p = 0.0532) than those with high *MUC4* expression (OS = 14.96; n= 35). Therefore, high *MUC1* expression and low *MUC4* expression negatively impact (p = 0.0344) OS in adult glioma patients (Fig. 3A).

**Fig. 3.**
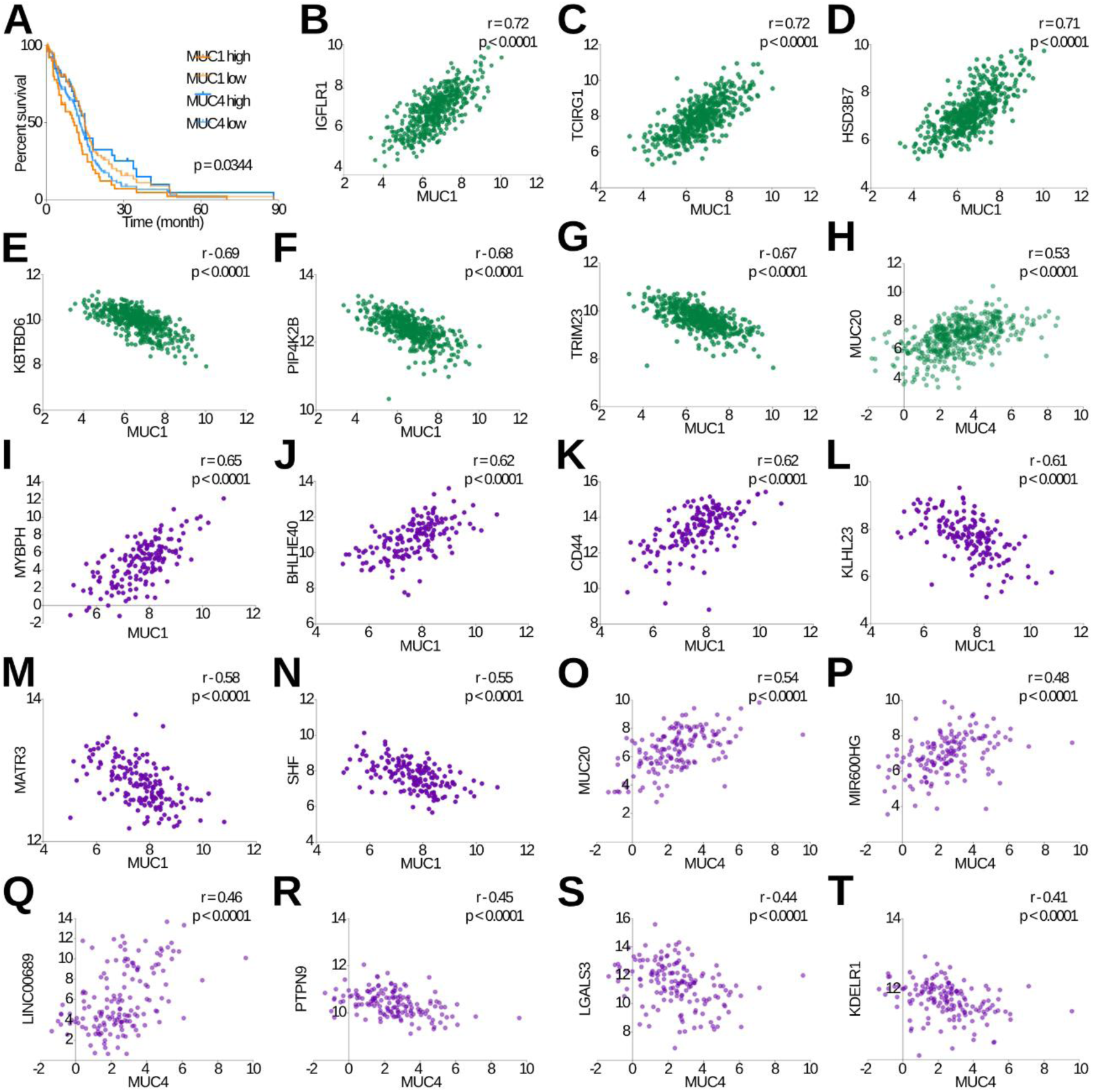
High *MUC1* expression and low *MUC4* expression predict poor OS and constitute a gene signature of coexpressed genes. (A) Kaplan‒Meier analysis of glioma patients (N = 153) revealed that high *MUC1* and low *MUC4* expression predicted worse OS. In the non-GBM cohort, genes coexpressed with MUC1 included *IGFLR1* (B), *TCIRG1* (C), *and HSD3B7* (D), whereas genes downregulated with MUC1 included *KBTBD6* (E), *PIP4K2B* (F) and *TRIM23* (G). For *MUC4*-related genes, strong coexpression was observed only for *MUC20* (H). In the GBM cohort, *MUC1* was coexpressed with genes such as *MYBPH* (I), *BHLHE40* (J) and *CD44* (K), while *KLHL23* (L), *MATR3* (M) and *SHF* (N) were downregulated. *MUC4* was coexpressed with *MUC20* (O), *MIR600HG* (P), and *LINC00689* (Q), while *PTPN9* (R), *LGALS3* (S) *and KDEL41* (T) were downregulated. D’Agostino-Pearson tests, Kaplan‒Meier curves and Spearman correlations were performed for statistical analysis. *MUC1* data are represented in dark colors, and *MUC4* data are represented in light colors. Green represents non-GBM or lower-grade glioma groups, and purple represents GBM data.

In addition, using data from cBioPortal, we explored genes coexpressed with *MUC1* and *MUC4* in gliomas. In the non-GBM patient cohort (N = 514), we found 1,757 genes with correlations above 0.40 and 1,114 with correlations below -0.40. The three genes that were most significantly (p < 0.0001) coexpressed with *MUC1* in non-GBM patients were *IGFLR1* (IGF-like family receptor 1; r = 0.72), *TCIRG1* (ATP6V0A3, ATPase H+ Transporting V0 Subunit A3; r = 0.72) and *HSD3B7* (hydroxy-delta-5-steroid dehydrogenase; r= 0.71) (Fig. 3B-3D). The three most negatively correlated genes were *KBTBD6* (Kelch Repeat and BTB Domain Containing 6; r -0.69), *PIP4K2B* (Phosphatidylinositol-5-Phosphate 4-Kinase Type 2 Beta; r -0.68) and *TRIM23* (Tripartite Motif Containing 23; r -0.67) (Fig. 3E-G). Regarding *MUC4*, *MUC20* was the unique gene with a strong positive correlation (r= 0.53; p <0.0001) (Fig. 3H), and no genes exhibited a strong negative correlation.

In the GBM patient cohort (N= 160), we identified 480 genes with correlations above 0.40 and 206 with correlations below -0.40 in relation to *MUC1* expression. The three genes most significantly (p <0.0001) positively coexpressed with *MUC1* were *MYBPH* (myosin binding protein H; r = 0.65), *BHLHE40* (basic helix-loop-helix family member E40; r= 0.62) and *CD44* (HCAM, homing cell adhesion molecule; r = 0.62) (Fig. 3I-K), while the most negatively coexpressed genes were *KLHL23* (Kelch Like Family Member 23; r -0.61), *MATR3* (Matrin 3; r -0.58) and *SHF* (Src Homology 2 Domain Containing F; r -0.55) (Fig. 3L-N). With respect to MUC4 expression, 77 genes had correlations above 0.40, and 7 had correlations below -0.40. The three genes most significantly (p <0.0001) positively coexpressed with *MUC4* were *MUC20* (Mucin 20; r =0.54), *MIR600HG* (MIR600 Host Gene; r =0.48) and *LINC00689* (Long Intergenic Non-Protein Coding RNA 689; r =0.46) (Fig. 3O‒Q), and the most negatively coexpressed genes were *PTPN9* (Protein Tyrosine Phosphatase Non-Receptor Type 9; r -0.45), *LGALS3* (Galectin 3; r -0.44) and *KDELR1* (KDEL Endoplasmic Reticulum Protein Retention Receptor 1; r -0.41) (Fig. 3R–T).

### 3.4 *MUC1* and coexpressed genes are related to innate and adaptive immune responses in gliomas, and *MUC4* and coexpressed genes are related to ion transport in GBM

Furthermore, to determine the biological role of genes coexpressed with *MUC1* and *MUC4* in gliomas, we conducted functional enrichment analysis. The 1,757 *MUC1* coexpressed genes in non-GBM patients were linked to innate immunity (cluster 1; ES= 31.05), antigen processing and presentation (cluster 2; ES= 11.37), and the regulation of cytotoxicity mediated by T and NK cells (cluster 3; ES= 3.36) (Fig. 4A). As the unique gene positively correlated with *MUC4* in non-GBM patients was *MUC20*, no enrichment analysis was possible. In GBM patients, the 480 genes coexpressed with *MUC1* were related to the innate immune response (cluster 1; ES= 6.51), positive regulation of proinflammatory factor production (cluster 2; ES= 5.39) and cell adhesion mediated by integrin and its signaling pathway (cluster 3; ES= 2.05) (Fig. 4B). In relation to *MUC4*, the 77 genes coexpressed in GBM patients were related to ion transport (cluster 1; ES= 2.07) (Fig. 4C).

**Fig. 4.**
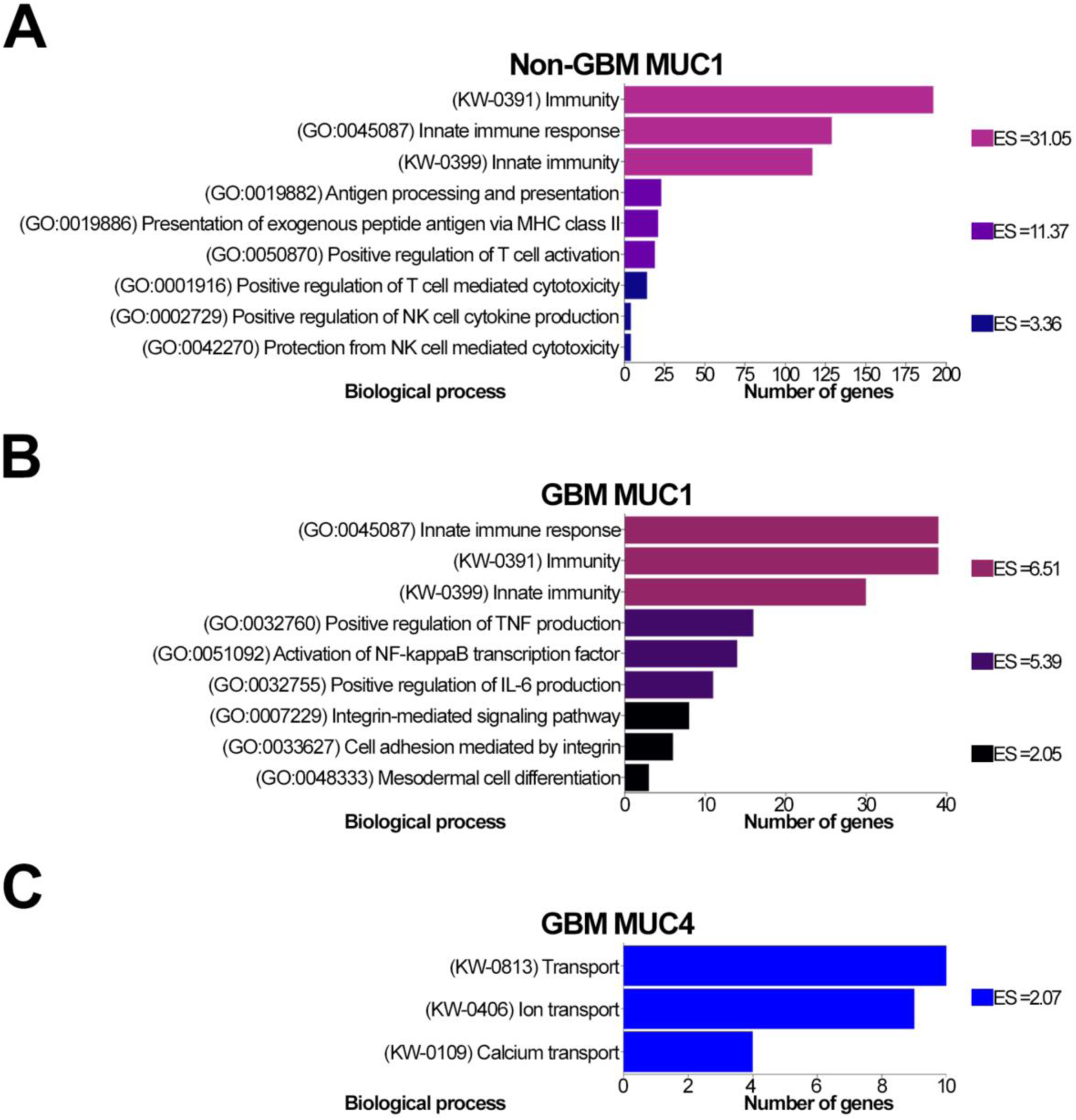
Genes correlated with *MUC1* and *MUC4* are linked to innate and adaptative immune responses, extracellular matrix-cell signaling and ionic transport in gliomas. Enrichment analysis of genes significantly positively correlated with *MUC1* and *MUC4* in gliomas was conducted using the DAVID bioinformatics resource. In the non-GBM cohort, the biological process of genes correlated with (A) *MUC1* involved the immune system (ES = 31.05), antigen processing and presentation (ES = 11.37), and regulation of cytotoxicity mediated by T and NK cells (ES = 3.36). In the GBM cohort, genes correlated with (B) *MUC1* were involved in the innate immune response (ES = 6.51), regulation of proinflammatory factors (ES= 5.39) and cell adhesion mediated by integrin (ES= 2.05). For (C) *MUC4*, correlated genes are involved in ion transport (ES = 2.07) in GBM. Statistical analysis was performed using the DAVID bioinformatics system. The enrichment score (ES) was used to rank the biological significance of each gene based on its p value.

### 3.5 The MUC1 SEA domain physically interacts with proteins from the innate and adaptive immune systems and an ECM receptor

Based on our previous enrichment analysis, we investigated the ability of the MUC1 protein to physically interact with key molecules in identified pathways using protein‒protein docking simulations. For this purpose, we selected RAGE, a critical receptor regulating innate immunity [59]; MHCII, essential for adaptive immunity [60]; and ITGA2, which mediates cell adhesion to the ECM and activation of protumor signaling pathways [61]. Due to the large size of the MUC1 protein, we focused on its SEA domain. As a control, we examined the interaction with Gal-3-BP, which is known to interact with MUC1 [57,58]. The interaction between the MUC1 SEA domain and Gal-3-BP had the lowest energy score of -793.1 (Fig. 5A), indicating a strong interaction. Additionally, RAGE exhibited the lowest energy score compared to that of the control, at -704.4 (Fig. 5B), followed by MHCII at -658.4 (Fig. 5C) and ITGA2 at -644.1 (Fig. 5D).

**Fig. 5.**
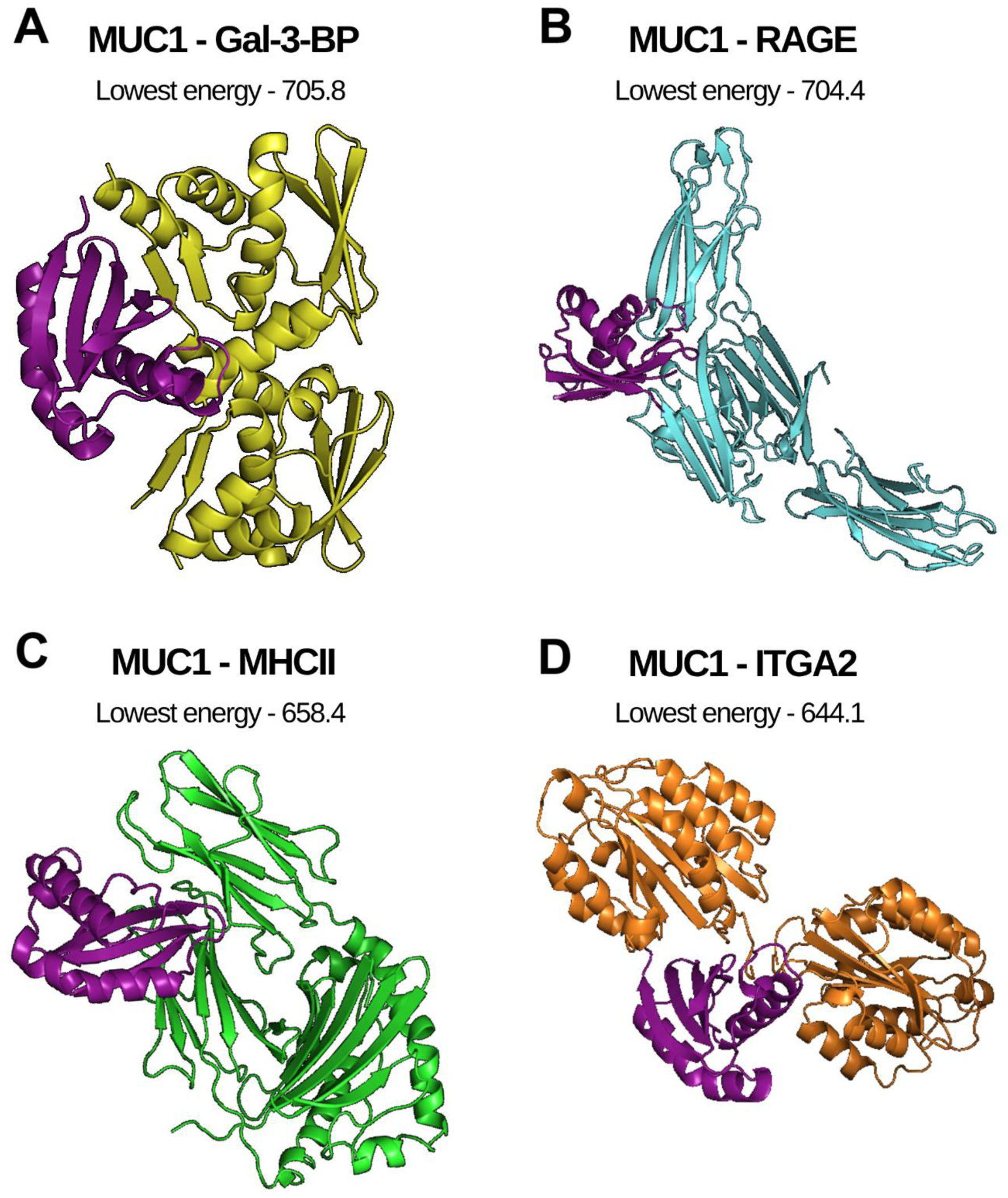
Structural models of MUC1 SEA domain interactions with immune response proteins and integrin alpha 2. Clustering analysis was conducted using ClusPro 2.0 to examine the interactions between the MUC1 SEA domain and key proteins of the innate and adaptive immune system, as well as ECM-related receptors. The interaction between Gal-3-BP and MUC1 served as a control (A). MUC1 demonstrated the ability to interact with RAGE (B), an innate immune protein; MHCII (C), a crucial molecule in the adaptive immune system; and ITGA2 (D), an adhesion receptor that mediates cell adhesion to the extracellular matrix and signaling. The MUC1 SEA domain is depicted in purple.

## 4. Discussion

Diffusely infiltrating gliomas are the most common primary CNS malignancies affecting the adult population and are classified into three distinct entities based on histopathology [8]. Characteristically, these neoplasms exhibit infiltrative growth, with tumor cells invading normal CNS tissues, thereby giving rise to recurrence and complicating conventional treatments [62,63]. Since the fifth edition of the WHO classification, the characterization of diffuse gliomas has heavily incorporated molecular alterations, which are crucial for defining both entity grades [64]. Consequently, molecular features have become increasingly important for glioma characterization, providing more accurate diagnoses and prognoses, reducing sampling errors and facilitating clinical decision-making [65].

Building upon the significance of molecular characterization in gliomas, GBM has emerged as the most severe and incurable cancer of the CNS, and current standard treatments are unable to address its heterogeneity, mutability and invasiveness [66]. Intra-and intertumor heterogeneity complicates the targeting of all tumor cells and patients receiving the same drug [67]. Therefore, novel biomarkers are urgently needed for diagnosis, personalized treatment, therapeutic response assessment, and relapse identification [68,69].

In the early 1980s, cancer antigen 125 (CA125), corresponding to the cleaved extracellular domain of MUC16, was routinely used as a serum biomarker for epithelial ovarian cancer [70,71], indicating the potential of mucins as diagnostic and therapeutic targets [72]. However, the role of mucins in gliomas, which are nonepithelial tumors, remains to be elucidated. Earlier studies from our group demonstrated the clinical significance of *MUC16* mutation [73] and *MUC17* mutation and methylation [74] on glioma patient survival. In the present study, through a retrospective analysis, we investigated for the first time the correlation of *MUC1* and *MUC4* expression with glioma malignancy and survival.

Epigenetic regulation of mucin promoters has been reported to influence prognosis and acquired resistance in non-small cell lung cancer [75], pancreatic ductal adenocarcinomas [76,77], and other epithelial tumors [78–80]. Using methylation data from the CGGA patient cohort (N= 99), we found that decreased *MUC1* methylation was significantly associated with increased glioma malignancy, while *MUC4* methylation showed the opposite pattern and was significantly increased in higher-grade gliomas. Thus, the *MUC1* and *MUC4* promoters act as epigenetic sensors in glioma, with *MUC1* being hypomethylated and *MUC4* being hypermethylated in GBM patients. In pancreatic ductal adenocarcinomas, *MUC1* and *MUC4* promoter hypomethylation predicts decreased OS and is associated with distant metastasis in stage IIA and IIB patients [76]. *MUC1* and *MUC4* mRNA expression is known to be regulated through promoter methylation at CpG sites [81–83]. In a cohort of glioma patients (N= 709) from the CGGA, we analyzed the expression of *MUC1* and *MUC4*, and the results were consistent with the methylation levels. *MUC1* was significantly more highly expressed in high-grade gliomas and GBM, while *MUC4* expression was lower in this cohort. Consistent with our results, KIM *et al*. (2020) [25] evaluated *MUC1* expression in paired normal brain (N = 70) and glioma (N = 30) samples with varying malignancy grades and found that *MUC1* upregulation was greater in gliomas than in normal tissues and was universally found in gliomas. However, MUC1 upregulation was statistically significant only in high-grade gliomas, such as GBM. Moreover, in the human GBM cell line U87, which is positive for epidermal growth factor receptor variant III (EGFRIII), *MUC1* stabilizes this receptor and confers resistance to temozolomide [26]. Taken together, these findings suggested that *MUC1* expression increases with glioma aggressiveness. In contrast to our findings, using formalin-fixed paraffin-embedded sections of glioma tumors (N=60) with different glioma grades, QUESNEL *et al.,* (2022) [28] reported increased immunohistochemistry (IHC) scores for *MUC4* and matrix metallopeptidase 9 in high-grade gliomas, and the expression rate was significantly greater in grade 4 gliomas. The increased methylation and decreased expression of *MUC4* found in our study may reflect the heterogeneous nature of gliomas, our larger cohort and our classification cohorts according to the WHO2021. Consistent with our results, in breast carcinogenesis, IHC staining of MUC4 in 26 normal breast tissues versus 298 aggressive breast tumors revealed that tumor progression was accompanied by decreased MUC4 expression, and in invasive breast carcinoma, *MUC4* promoter hypermethylation and MUC4 downregulation were correlated with increased numbers of tumor-infiltrating immune cells [84]. The combined evaluation of mucin expression is an effective strategy for detecting prognostic biomarkers and MUC4 should be further explored in gliomas [85].

To confirm that *MUC1* and *MUC4* methylation affects their expression in gliomas, we established a new patient cohort using TCGA data deposited on cBioPortal, where we simultaneously assessed the methylation and expression of these genes. In both the non-GBM (N = 530) and GBM (N = 137) patient cohorts, overall increased methylation of these genes significantly (p < 0.0001) decreased their expression. However, in GBM cohort, increased MUC4 methylation did not significantly correlate with decreased MUC4 expression, but there was a tendency (p = 0.1344). Thus, the DNA methylation status of *MUC1* and *MUC4* correlates with their expression and is a potential marker of malignancy in gliomas. Furthermore, our study demonstrated that *MUC1* and *MUC4* methylation are closely correlated in gliomas but in different directions. In the non-GBM cohort, *MUC1* and *MUC4* methylation levels tended to increase. However, in the GBM cohort, an increase in *MUC4* methylation led to a decrease in *MUC1* methylation. To our knowledge, this is the first time that *MUC1* and *MUC4* methylation levels have been correlated with each other in tumors in general. Regarding the expression of these mucins per se, a significant (p < 0.0001) positive correlation between *MUC1* and *MUC4* expression was identified by KAMIKAWA *et al.,* (2015) [86] in oral squamous cell carcinoma samples (N = 206), with double-positive patients demonstrating worse OS. Thus, the underlying epigenetic mechanism regulating these genes in gliomas might be relevant as a molecular marker and therapeutic target.

Furthermore, in the survival analysis, we demonstrated that *MUC1* and *MUC4* expression levels significantly (p = 0.0344) impacted the OS of glioma patients (N = 153), as patients with high *MUC1* expression and low *MUC4* expression had a worse prognosis. We hypothesize that high *MUC1* expression in high-grade gliomas may compensate for *MUC4* downregulation in glioma aggressiveness [87].

Our study also revealed genes that were significantly (p < 0.0001) coexpressed and downregulated with *MUC1* and *MUC4* in gliomas. Among the genes coexpressed with *MUC1* in the non-GBM cohort, *TCIRG1* and *HSD3B7* have been described in relation to tumor-associated immune system functions and as biomarkers in gliomas [88–90]. The role of *IGFLR1* in gliomas needs to be elucidated; however, in clear cell renal cell cancer, *IGFLR1* promoter methylation decreases with pathological stage progression, and increased *IGFLR1* expression is associated with poor OS and immune infiltration [91]. Moreover, high *IGFLR1* expression was observed in specific T-cell subsets (CXCL13^+^BHLHE40^+^ TH1-like cells and CD8^+^ exhausted T cells) in colorectal cancer [92]. The roles of genes downregulated with *MUC1* expression (*TRIM23, PIP4K2B* and *KBTBD6*) in gliomas remain to be elucidated. In breast tumors, low *PIP4K2B* expression was associated with increased tumor size and distant metastasis [93]. Furthermore, *KBTBD6* expression was linked to treatment response in pituitary adenoma [94].

Among the genes coexpressed with *MUC1* in GBM patients, *MYBPH* and *CD44* have been previously described as biomarkers in gliomas, predicting poor prognosis [95–97]. In a bioinformatics analysis, QI *et al.,*(2023)[98] reported that *BHLHE40* was overexpressed in 7 of 33 tumor tissues, including GBM tissues, and predicted worse OS in both high- and low-grade gliomas. Among the genes downregulated with *MUC1* expression, *SHF* plays a tumor suppressor role in GBM by negatively regulating signal transducer and activator of transcription 3 (STAT3) activity, and its low expression predicts worse OS [99]. Additionally, low *KLHL23* and *MATR3* expression is also associated with poor survival in patients with different tumors [100–103].

In the GBM cohort, the genes coexpressed with *MUC4* are noncoding RNAs with varying roles in gliomas. ZHU *et al.,* (2020a)[104] reported that high *MIR600HG* expression is associated with improved outcomes in glioma patients. Conversely, high expression of *LINC00689* plays an oncogenic role and predicts poor OS [105,106]. Additionally, among the genes downregulated with *MUC4* expression in GBM, *PTPN9* is a tumor suppressor gene in glioma, and its overexpression reduces glioma cell proliferation [107]. Moreover, *KDELR1* and *LGALS3* overexpression are linked to malignancy and poor prognosis [108–110].

Interestingly, we found that *MUC4* expression is positively correlated with *MUC20* expression in both non-GBM and GBM gliomas. *MUC20* is a transmembrane mucin located on chromosome locus 3q29 close to *MUC4* [18]. The functional importance of *MUC20* has been demonstrated in several cancers [111–113], but its role in gliomas remains to be further elucidated. In a similar study on different tumors, JONCKHEERE and VAN SEUNINGEN (2018)[114], through bioinformatics analysis, evaluated genes correlated with *MUC4* expression in epithelial cancers and found a close correlation between *MUC16* and *MUC20*. Their study showed that the *MUC4/MUC16/MUC20* expression signature predicted worse OS in pancreatic, colon and stomach cancers. The prognostic role of *MUC16* mutations was previously described by our group [73]; however, the *MUC4/MUC20* axis is still unexplored in gliomas and could present prognostic value.

Taken together, our study revealed that genetic signatures related to *MUC1* and *MUC4* expression, such as immune cell infiltration, cell motility, tumor growth, and poor OS, are correlated with genes related to tumor aggressiveness. Moreover, we identified genes whose functions in gliomas have not yet been explored, highlighting new targets for glioma research, especially the *MUC4*/*MUC20* axis.

We conducted an enrichment analysis of the genes coexpressed with *MUC1* and *MUC4* in gliomas. In both non-GBM and GBM gliomas, *MUC1*-correlated genes were primarily involved in the immune response. GBM is highly infiltrated by immune cells, including microglia, monocyte-derived macrophages, and myeloid-derived suppressor cells, and there is a positive correlation between the accumulation of these cells and glioma grade [115,116]. Additionally, some studies have concluded that natural killer (NK) cells also infiltrate brain tumors [117]. In the early phases of tumor development, antigen-presenting cells and even tumor cells activate specific cytotoxic T cells through the major histocompatibility complex to impair tumor growth [118]. However, during tumor progression, GBM becomes immunosuppressive, leading to immune tolerance and tumor growth [119]. The gene signature found in your study likely represents a population of immune infiltrating cells.

In addition to our enrichment analysis of MUC1-related genes in GBM, our study also highlighted their role in mediating cell-matrix adhesion. The migration and invasion of glioma cells into healthy brain parenchyma necessitate the modification of ECM components and the expression of receptors that mechanically link tumor cells with the ECM [12]. Integrins are the main family of receptors that mediate cation-dependent cell adhesion to ECM components [120]. Activation of integrins initiates downstream signaling pathways that regulate cell migration, invasion, proliferation and survival [121]. In GBM, specific integrin members (*ITGA2*, *ITGA3*, *ITGA5* and *ITGB1*) are overexpressed, which is correlated with poorer OS [122,123]. These findings emphasize the role of MUC1-correlated genes in immune response modulation and maintenance of tumor aggressiveness through ECM interactions.

Furthermore, we found that genes coexpressed with *MUC4* in GBM patients are associated with ion transport. In gliomas, the regulation of ion channels is essential for gene expression, cell migration and proliferation [124]. The direct involvement of *MUC4* in the regulation of these channels has yet to be explored in tumors. Therefore, we propose that *MUC1* and *MUC4* have distinct roles in gliomas, and this could be further explored in these tumors.

Finally, we explored the potential of the MUC1 SEA domain to physically interact with several key proteins that are involved in signaling pathways through our enrichment analysis. MUC1 is known to be a natural ligand of galectin-3 that supports tumor development [58,125,126], and Gal-3BP is necessary for interaction with galectin-3, which has been identified as a cancer- and metastasis-associated protein [127] that plays a role in the innate immune response [57]. In our study, through computational modeling, compared with the pattern of interaction between MUC1 and Gal-3BP, we determined that the MUC1 SEA domain can form complexes with RAGE, a pivotal transmembrane receptor in the innate immune response [128,129]; MHCII, involved in antigen presentation and the development of functional adaptive immunity [60]; and ITGA2, which mediates cell-ECM contact and activates integrin signaling pathways [130,131]. These interactions with MUC1 in glioma warrant further elucidation and may be significant signaling discoveries, which are likely involved in the aggressiveness of these tumors.

## 5. Conclusions

In summary, for the first time, this study demonstrated that *MUC1* and *MUC4* are differentially methylated in adult diffuse gliomas and are correlated with tumor grade and glioma subtype. In non-GBM gliomas, these genes exhibit increased methylation together, while in GBM, increased *MUC4* methylation decreases *MUC1* methylation. We verified that the expression of these genes is correlated with the methylation level of their promoters. Furthermore, in this study, we identified genetic signatures of genes coexpressed or downregulated with *MUC1* and *MUC4* in diffuse gliomas. We identified new potential genes, such as *MUC1* and *MUC4*, and coexpressed genes (for example, *IGFLR1, TRIM23, PIP4K2B* and *KBTBD6)* that remain to be elucidated in these tumors. Specifically, we identified a conserved axis in gliomas involving *MUC4* and *MUC20* coexpression and demonstrated that *MUC1* coexpressed genes are involved mainly in the immunological response in gliomas, whereas *MUC4* coexpressed genes are involved in ion transport in GBM. Finally, we demonstrated that the MUC1 SEA domain may physically interact with the receptors RAGE, MHCII and ITGA2, which are membrane proteins related to the immune response, tumor infiltration and aggressiveness.

## Data Availability

All data produced in the present study are available upon reasonable request to the authors

## Authors contributions

G.M and V.F. screened and taken responsibility for the integrity of the data and the accuracy of the data analysis, performed bioinformatic analysis, wrote and edited the manuscript. V.F also supervised the work and acquired funding.

## Data availability

The data generated in this study are available within the article. Raw data will be available upon request.

